# COVID-19 Outbreak Control Strategies and their Impact on the Provision of Essential Health Services in Ghana: An Explanatory-Sequential Study

**DOI:** 10.1101/2022.12.12.22283342

**Authors:** Duah Dwomoh, Isaac Yeboah, Rawlance Ndejjo, Steven Ndugwa Kabwama, Justice Moses Aheto, Anne Liu, Siobhan Lazenby, Fidelia Ohemeng, Sylvia Akpene Takyi, Ibrahim Issah, Serwaa Akoto Bawuah, Rhoda K. Wanyenze, Julius Fobil

## Abstract

**Background:** The COVID-19 pandemic has led to substantial interruptions in critical health services, with 90% of countries reporting interruptions in routine vaccinations, maternal health care and chronic disease management. The use of non-pharmaceutical interventions (NPIs) such as lockdowns and self-isolation had implications on the provision of essential health services (EHS). We investigated exemplary COVID-19 outbreak control strategies and explored the extent to which the adoption of these NPIs affected the provision of EHS including immunization coverage and facility-based deliveries. Finally, we document core health system strategies and practices adopted to maintain EHS during the early phase of the pandemic.

**Methods:** This study used an explanatory sequential study design. First, we utilized data from routine health management information systems to quantify the impact of the pandemic on the provision of EHS using interrupted time series models. Second, we explored exemplary strategies and health system initiatives that were adopted to prevent the spread of COVID-19 infections while maintaining the provision of EHS using in-depth interviews with key informants including policymakers and healthcare providers.

**Results:** The COVID-19 pandemic and the interventions that were implemented disrupted the provision of EHS. In the first month of the COVID-19 pandemic, Oral Polio and pentavalent vaccination coverage reduced by 15.2% [95% CI = −22.61, −7.87, p<0.001] and 12.4% [95% CI = 17.68, −7.13; p<0.001] respectively. The exemplary strategies adopted in maintaining the provision of EHS while also responding to the spread of infections include the development of new policy guidelines that were disseminated with modified service delivery models, new treatment and prevention guidelines, healthcare workforce capacity building on outbreak control strategies, the use of telemedicine and medical drones to provide EHS and facilitate rapid testing of suspected cases.

**Conclusion:** The implementation of different NPIs during the peak phase of the pandemic disrupted the provision of EHS. However, the Ministry of Health leveraged the resilient health system and deployed efficient, all-inclusive, and integrated infectious disease management and infection prevention control strategies to maintain the provision of EHS while responding to the spread of infections.

## Introduction

The severe acute respiratory syndrome coronavirus 2 (SARS-CoV-2), the coronavirus that causes coronavirus disease 2019 (COVID-19) led to an unprecedented number of deaths and morbidities across the globe [1]. Ghana recorded its index COVID-19 case on the 12th of March 2020 and as of the 20^th^ of October 2022, the country has recorded a total of 170321 cases and 1460 COVID-19-induced deaths. Governments across the globe implemented several non-pharmaceutical interventions (NPIs) and vaccination strategies to reduce the burden of the pandemic on health systems and prevent new infections and deaths and increase access to essential health services (EHS). EHS encompass services with high priority guided by a country’s health system and burden of disease aimed at preventing communicable diseases, reducing maternal mortality and child morbidity, preventing acute chronic conditions, and managing emergency conditions. It focuses on high-priority service delivery as listed in the WHO guidance and tools manual. It includes prevention of communicable diseases, services related to reproductive health (care during pregnancy and childbirth), and care of vulnerable populations such as infants and adults. The high-priority service delivery also includes the provision of medications, continuity of critical inpatient therapies, and auxiliary services such as laboratory services and blood bank services to patients. These NPIs disrupted EHS and compounded the social-economic effects on the populace which made them unsustainable in the long term [2]. As COVID-19 spread, the provision of EHS was negatively affected due to weak health systems, limited-health service infrastructure and poor health financing systems [3]. EHS are disrupted because of the unavailability of health workers, infection of health workers and shortage of essential drugs [4, 5]. Other supply dynamics that contribute to the decline in the use of EHS are lockdown policies and stay-at-home orders [6, 7]. The clients’ fear of contracting the infectious disease results in a decrease in the use of EHS [8]. Understanding how a broader range of NPIs strategies have been instituted to mitigate against the spread of COVID-19, as well as any unexpected adverse effects on EHS demand is essential for future health system planning and pandemic preparedness. Few studies have investigated infection prevention strategies, the impact of COVID-19 on EHS and exemplary interventions implemented to improve the uptake of EHS during the peak phase of the pandemic. We provide evidence of the effect of the pandemic, identify and highlight the best practices adopted by Ghana in maintaining the provision of EHS while also responding to the COVID-19 pandemic using different infection prevention control strategies. The present study is underpinned by the statistical and socio-ecological models that evaluates health systems’ resilience to fight the pandemic vis a vis the implementation of policies to increase the use and delivery of EHS.

## Methods

This study used an explanatory sequential design in which we collected and analyzed time series data on EHS and then followed the results up with a qualitative phase of in-depth interviews with key informants among healthcare workers and policymakers to explore mitigation strategies adopted to curb the spread of infection and maintain the provision of EHS.

### Study sites

The study was conducted across the 16 administrative regions of Ghana between March 2020 to December 2021. However, the secondary data analysis of EHS started in January 2018 to December 2021 for all health facilities that report monthly data to routine health management information system.

## Quantitative study design

### Data source for interrupted time series analysis

This was a retrospective ecological time series study design using routine time series data on EHS between January 2018 and December 2021. The EHS data was a continuous sequence of observations at the district level taken repeatedly over equal monthly intervals. Interrupted Time Series (ITS) is a quasi-experimental study design with a potentially high degree of internal validity [9]. In this ITS study, a time series of the EHS outcomes of interest were used to establish an underlying trend in EHS utilization, which was assumed to be interrupted by the COVID-19 pandemic on the 12^th^ of March 2020. The ITS was used to model a hypothetical counterfactual scenario under which we assume that had COVID-19 not taken place, the trend on the provision and use of EHS would have continued unchanged (that is: the expected trend, in the absence of COVID, given the pre-existing trend). This counterfactual scenario provides a comparison for the evaluation of the impact of COVID-19 on EHS by examining any change occurring in the period after the first cases of COVID-19 were recorded in Ghana on March 12, 2020. We hypothesized a gradual effect of COVID-19 on EHS (that is, one month lag period before any observed effect).

### Outcome measures

The study analyzed eight EHS indicators: Antenatal care attendance, facility delivery, post-natal care attendance, oral polio vaccination, Bacille Calmette-Guerin (BCG) vaccination, Measles-Rubella vaccination, and outpatients’ visits per 1000 populations (Table 1).

**Table 1:** Outcome indicator definitions

| Indicator | Definition | Numerator | Denominator | Source of denominator |
| --- | --- | --- | --- | --- |
| BCG Coverage | The proportion of children under 1 year receiving BCG vaccine | Number of children under 1 year receiving the BCG vaccine in the period | Number of children under 1 year (estimated as 4% of the population) | Ghana Statistical Service from the 2010 Population and Housing Census |
| Oral Polio vaccination | The proportion of children under 1-year receiving oral polio (OPV1) vaccine | Number of children under 1 year receiving the OPV1 vaccine in the period | Number of children under 1 year (estimated as 4% of the population) | Ghana Statistical Service from the 2010 Population and Housing Census |
| Pentavalent vaccination | The proportion of children under 1 year receiving Pentavalent vaccine | Number of children under 1 year receiving the Penta 1 vaccine in the period | Number of children under 1 year (estimated as 4% of the population) | Ghana Statistical Service from the 2010 Population and Housing Census |
| Measles-Rubella Coverage | The proportion of children under 1 year receiving Measles-Rubella Vaccine | Number of children under 1 year receiving the Measles-Rubella vaccine in the period | Number of children under 1 year (estimated as 4% of the population) | Ghana Statistical Service from the 2010 Population and Housing Census |
| Antenatal Care Coverage | The proportion of pregnant women receiving antenatal care during pregnancy (at least once). | Total number of antenatal registrants in a specified period | Total number of expected pregnancies of the catchment area within the specified period | Ghana Statistical Service from the 2010 Population and Housing Census |
| Skilled delivery | Percentage of deliveries conducted by skilled attendants (nurses and doctors). | The number of deliveries supervised by doctors or nurses in the specified period. | Number of expected pregnancies (estimated as 4% of the population) | Ghana Statistical Service from the 2010 Population and Housing Census |
| Post-natal coverage | The proportion of PNC registrants seen after delivery | Number of PNC registrants (within 48 hours) | Number of expected pregnancies (estimated as 4% of the population) | Ghana Statistical Service from the 2010 Population and Housing Census |
| OPD attendance per 1000 population | The rate of facility visits per 1000 population | The number of OPD attendance per month for the index year | Total population per month for the index year | Ghana Statistical Service from the 2010 Population and Housing Census |

Source: District Health Management Information System: Ghana Health Service

### Statistical Analysis

Descriptive analysis using mean, standard deviation, minimum and maximum observations, median and interquartile range, and tools from time series were used to explore the distribution of the outcome measures and identified the underlying trend, seasonal patterns, and outliers in the EHS data. We tested the normality of all coverage indicators using the Jarque-Bera skewness-kurtosis test (Table 2).

**Table 2:**
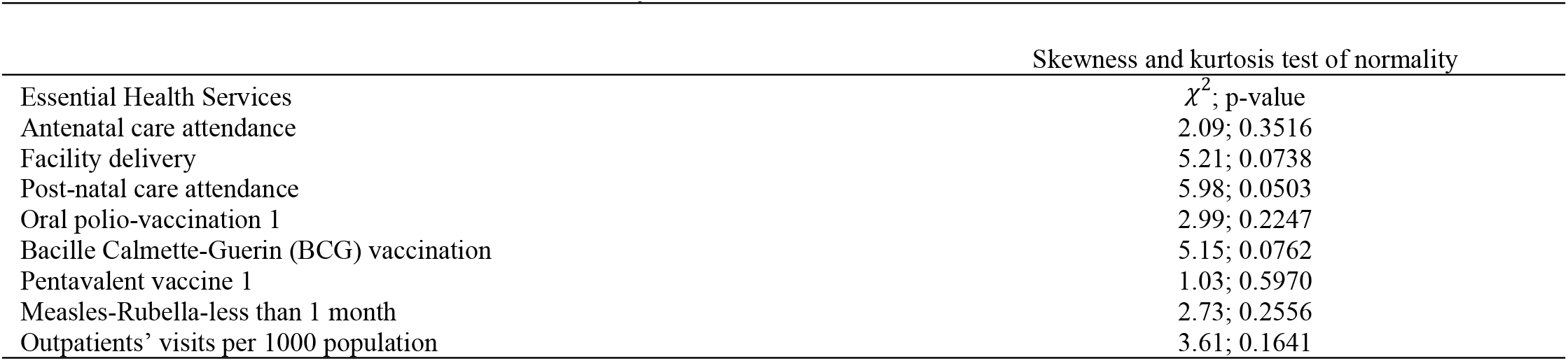
Skewness and kurtosis test of normality

Our ITS analysis involved three different statistical models. These models were fitted to EHS data before and during COVID. We fitted ordinary least square regression models to quantify the impact of COVID-19 on the provision of EHS. We used the Fourier terms (pairs of sine and cosine functions) to adjust for seasonality and other long-term trends (Bhaskaran et al. 2013). The following segmented harmonic ITS regression model with Newey West standard errors that adjusts for seasonality in EHS time-series dependent outcomes with sine/cosine pair was used:

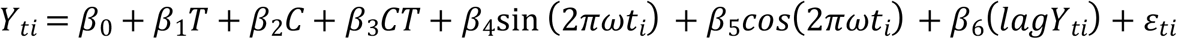

*Y*_*ti*_ is the coverage of EHS in the *t*th month of the *i*th year; *t* values range from 1 to 12; *i* values range from 1 to *K*, where *K* is the number of years under observation (2018-2021). *C* is a dummy variable indicating observation collected before COVID (C=0) or after (C=1). *T* is a continuous variable that indicates the time in months passed from the start of the observational period (2018-2021).

*β*_0_ represents the baseline level of EHS at *T* = 0, *β*_1_ is interpreted as the change in EHS associated with a unit increase in time and it represents the underlying pre-COVID-19 trend), *β*_2_ is the level change in EHS after the index case of COVID-19 was recorded on 12^th^ March 2020 and *β*_3_ indicates the slope change following the COVID-19 pandemic. *ε*_*ti*_ are independently and identically distributed normal random variables with *E*[*ε*_*ti*_] = 0 and *Var*[*ε*_*ti*_] = σ^2^. ω reflects the period within a year timespan and in our case, 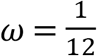 to reflect monthly data. Our proposed model describes the seasonal behaviour of EHS by both sine and cosine functions with symmetric rise and fall over a period of a full year. For both the Generalized Poisson and the negative binomial regression models, which are required for modeling under and over-dispersed EHS outcomes respectively, we included total OPD cases as an offset variable to convert the EHS outcomes into a rate and adjust for any potential changes in the OPD attendance over time. We estimated the counterfactual by assuming that no COVID-19 case was recorded in Ghana and there was no immediate nor sustained effect of the pandemic. We graphically assessed autocorrelation by examining the plot of residuals and the partial autocorrelation function. This was followed by the Cumby–Huizinga (Breusch-Godfrey) general test for autocorrelation [10]. We employed the Newey-West method [11] to generate more robust standard errors that were valid even when there is the presence of heteroscedasticity and autocorrelation.

### Data sources and Geospatial methods for regional distribution of cumulative confirmed cases

We obtained the number of cumulative confirmed COVID-19 cases from the Ghana Health Service website (https://www.ghs.gov.gh/covid19/archive.php) for the period of 12th March 2020 to 31st December 2021. The regional shapefile for Ghana was downloaded from GADM (https://gadm.org/download_country.html). The rgdal, leaflet and tmap packages in *R* software version 4.1.1 and RStudio were used for Geospatial data preparation [12].

### Qualitative Study Design

We used the socio-ecological framework for understanding infection prevention control strategies and measures adopted to maintain EHS and a phenomenological approach to guide this study. The socio-ecological framework describes population-based interventions at individual, interpersonal, institutional, community and public policy levels that influence sustaining essential health services during Covid-19 in Ghana. The phenomenological approach to this study is based on the premise that disrupted EHS, barriers and strategies to maintaining EHS and infection prevention control is best understood from the experiences of frontline health workers, policymakers and implementers of policies and programs. This descriptive exploratory study had a qualitative design using a semi-structured interview guide. The study features Interpretative Phenomenological Analysis (IPA) and narrative approaches. Frontline health workers shared their lived experiences with how they controlled the spread of infections while providing EHS. On the other hand, implementers and policymakers shared their experiences on policy initiatives and knowledge about how they reduce the spread of infections while maintaining EHS using the narrative approach. A thematic analysis was used as a tool to identify key themes emerging from the interviews. A deductive approach was chosen to better focus on the experiences and perspectives of frontline healthcare providers and policymakers.

### Participants recruitment

Individuals were considered eligible if they were frontline health workers, implementers, policymakers and stakeholders who worked during the COVID-19 pandemic in regions with high recorded COVID-19 cases. Participants were recruited using purposive sampling based on the responsibilities that were assigned to these individuals during the peak phase of the pandemic. The study purposively selected healthcare providers, policymakers, stakeholders and program implementers. For clarity, health workers who worked in health facilities during the early phase of the COVID-19 pandemic were referred to as frontline health workers and were recruited. Policymakers, implementers and stakeholders who were directly involved with decision-making at the community, district, regional and national levels regarding COVID-19 were also recruited. Recruitment took place between March and May in the Greater Accra Region of Ghana. Potential participants were asked if interested in participating in the study before interviews were conducted. Informed consent was obtained before the interview was conducted by the research assistants. A total of 32 participants (20 frontline health workers and 12 policymakers, implementers, and stakeholders) were involved in this study.

### Data Collection

The purpose and significance of the study were communicated to the study participants and interviews were scheduled at the convenience of the study participants. Frontline health workers were recruited from primary, secondary, tertiary, and quaternary health facilities. Policymakers, implementers, and stakeholders were also recruited from the national COVID-19 taskforce, regional rapid response team, municipal/district rapid response team, Ghana Health Service, Municipal health directorate and district health directorate. Two different interview guides were prepared before conducting interviews: one each for frontline health workers and policymakers, implementers and stakeholders. The interview guide for frontline health workers consists of contents such as services disrupted, challenges to the utilization of EHS and facility strategies to provide EHS. The interview guide for policymakers, implementers and stakeholders elicited information on disruptions of EHS, challenges to maintaining/restarting EHS and health system interventions to maintaining or restarting EHS. All the interview guides were in English. In addition, all interviews were conducted in English with most of the interviews at the office of the selected participants. Physical distance was maintained during face-to-face interviews while both the research assistant and research participant wore a face mask. All interviews were done on a face-to-face basis. Interviews lasted one hour to one hour and 30 minutes. After the interview, research assistants provided key points of the interview to validate the data as a form of checking. Research assistants wrote field notes on daily basis. Interviews were shared with authors to review and provide feedback. The daily feedback was done until saturation was reached.

### Qualitative data analysis

All interviews were audio-recorded, transcribed verbatim, exported to NVivo (Version 10) and analyzed using the six steps of thematic analysis described by Braun & Clarke [13]. The six steps of thematic analysis involve (1) data familiarization, (2) generating initial codes (3) identifying preliminary themes (4) reviewing themes (5) defining and naming themes; (6) reporting. First, the entire transcripts were read for an overall understanding of the data. Secondly, codes addressing research questions were identified. Thirdly, the initial codes identified were synthesized into wider themes. The themes from frontline health workers and policymakers and implementers were reviewed and synthesized. In the fifth stage, we drew up figures for thematic analysis where data for each theme were collected from all interview transcripts.

### Ethical considerations

The study was approved by the Ghana Health Service Ethical Review Committee (GHS-ERC) with a unique approval number GHS-ERC: 008/11/12.

## Results

### Geospatial distribution of cumulative confirmed cases by region

Our results indicate substantial regional variations in the number of cumulative confirmed cases in the country from 12th March 2020 to 31^st^ December 2021. The Greater Accra region recorded the highest number of cases (81932) (Figure 1).

**Figure 1.**
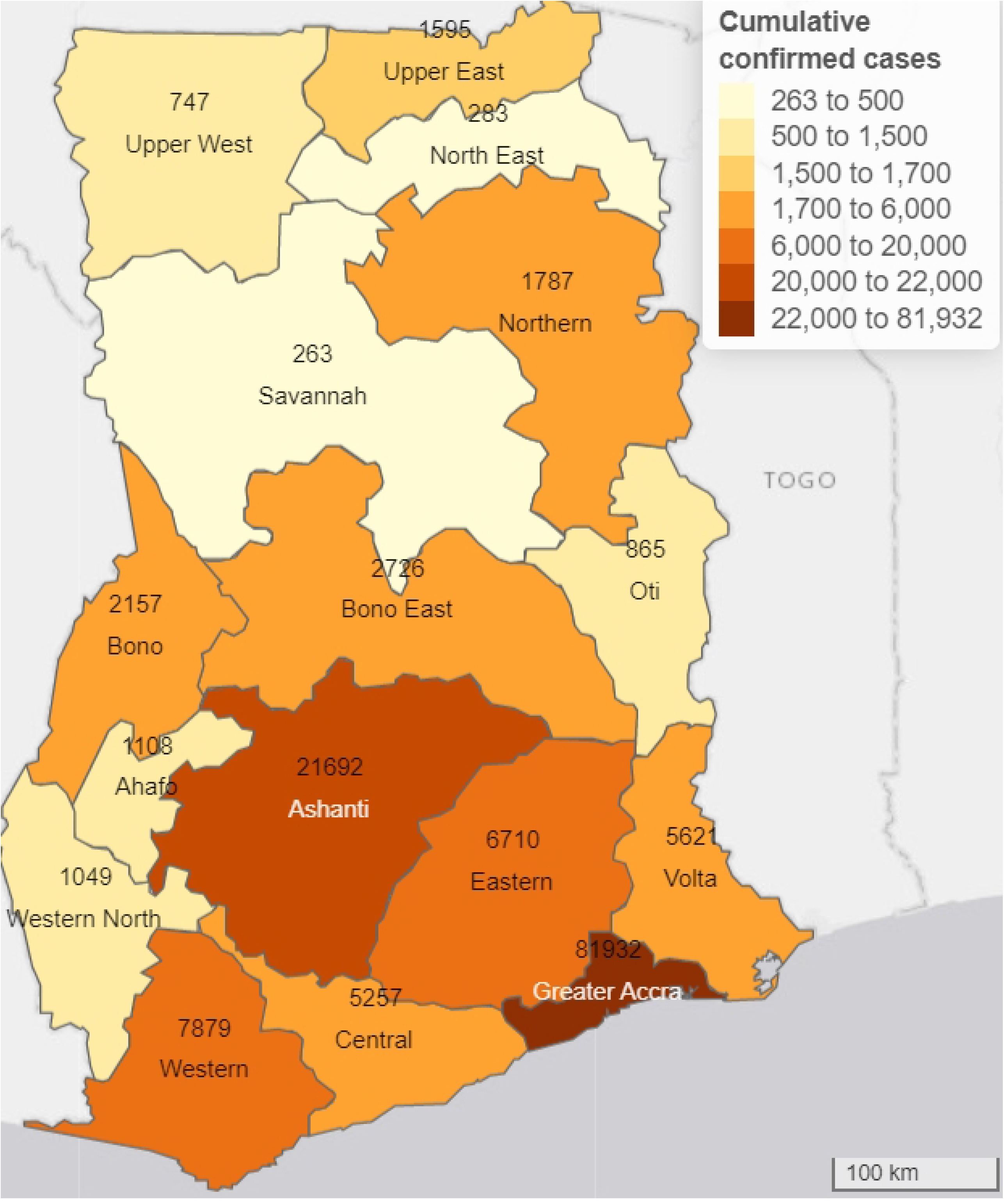
Geospatial distribution of cumulative confirmed cases of COVID-19 by regions in Ghana (12^th^ March 2020 to 31^st^ December 2021).

### Impact of COVID-19 on essential health service delivery

Table 3 shows the impact of COVID-19 on EHS delivery in Ghana. The pandemic had an immediate effect on BCG, OPV, Measles and Pentavalent vaccination as the coverages of these indicators declined within one month after Ghana recorded its first two index cases. However, it was observed that the pandemic had an immediate negative effect on the provision of EHS (OPV and Pentavalent vaccination coverages) as they declined within the first month of the pandemic. However, the decline in the provision of EHS was not sustained as utilization of EHS began to increase after the first few months of the pandemic. In the years prior to Ghana recording the first two index cases (1^st^ January 2018 to 11^th^ March 2020), OPV coverage was estimated at 88.3% [95% CI: 85.67, 91.26]. In the first month of the COVID-19 pandemic, OPV coverage decreased by 15.2% [95% CI = −22.61, −7.87, p<0.001] which corresponds to the immediate effect of the pandemic. However, this decline was not sustained as the monthly trend of OPV coverage relative to the pre-COVID trend increased by 1.04% on average per month [95% CI: 0.62, 1.46, p<0.001]. Our results on the post-COVID trend analysis showed that after 12^th^ March 2020, OPV coverage increased monthly at a rate of 1.2% [95% CI: 0.80, 1.50; p<0.001]. In the first month of the COVID-19 pandemic, Pentavalent vaccination coverage decreased by 12.4% [95% CI = 17.68, − 7.13; p<0.001] which corresponds to the immediate effect of the pandemic. Like OPV, this reduction in coverage was not sustained as the monthly trend of Pentavalent vaccination coverage increased thereafter relative to the pre-COVID trend by 0.9% on average per month [95% CI: 0.53, 1.22; p<0.001]. Our results on the post-COVID trend analysis showed that after 12^th^ March 2020, Pentavalent vaccination coverage increased monthly at a rate of 1.0% [95% CI: 0.72, 1.21%; p<0.001]. PNC attendance decreased immediately after the country recorded the first two index cases, but general OPD attendance has increased since the inception of COVID-19 in Ghana (Figure 2).

**Table 3:**
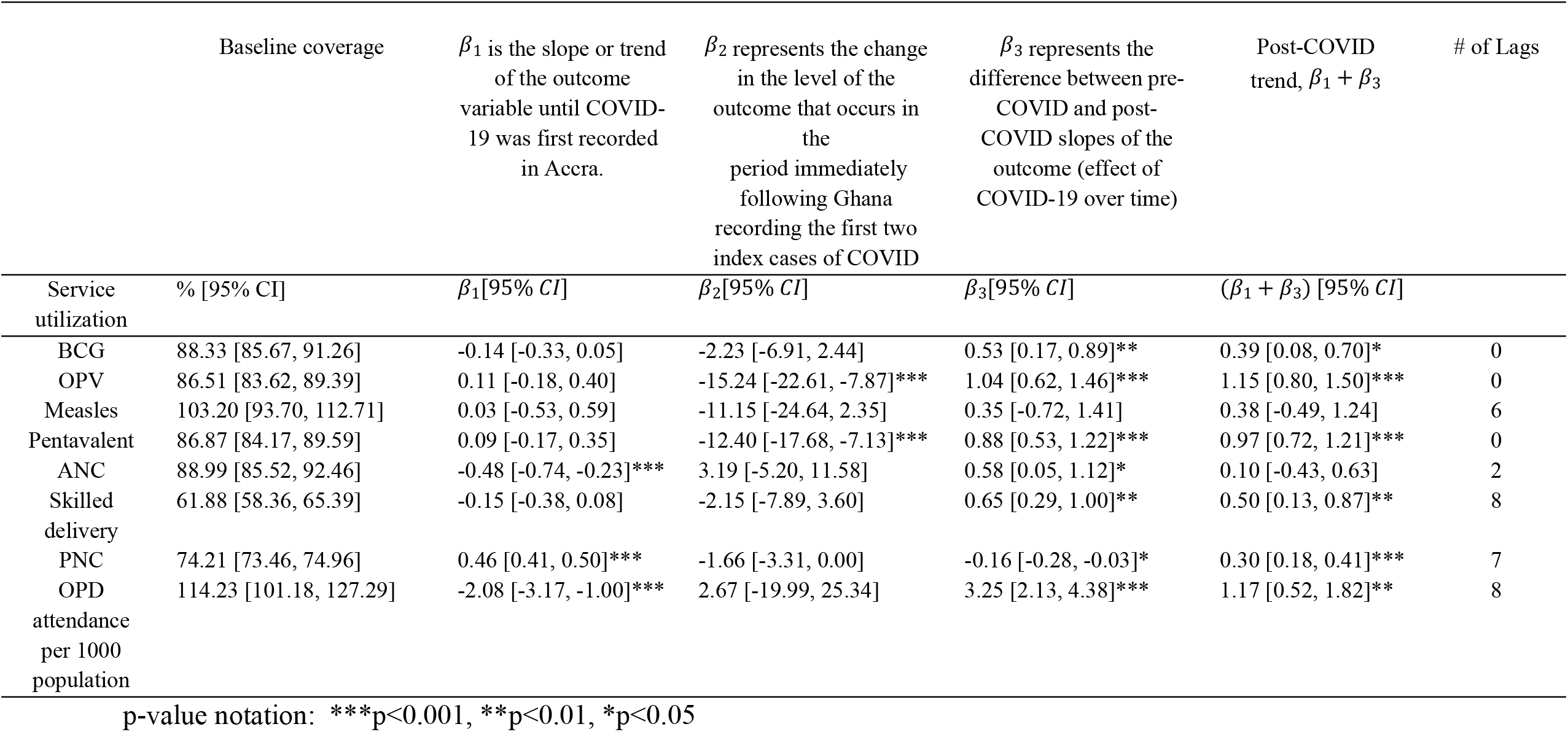
Impact of COVID-19 on essential health service delivery

**Figure 2.**
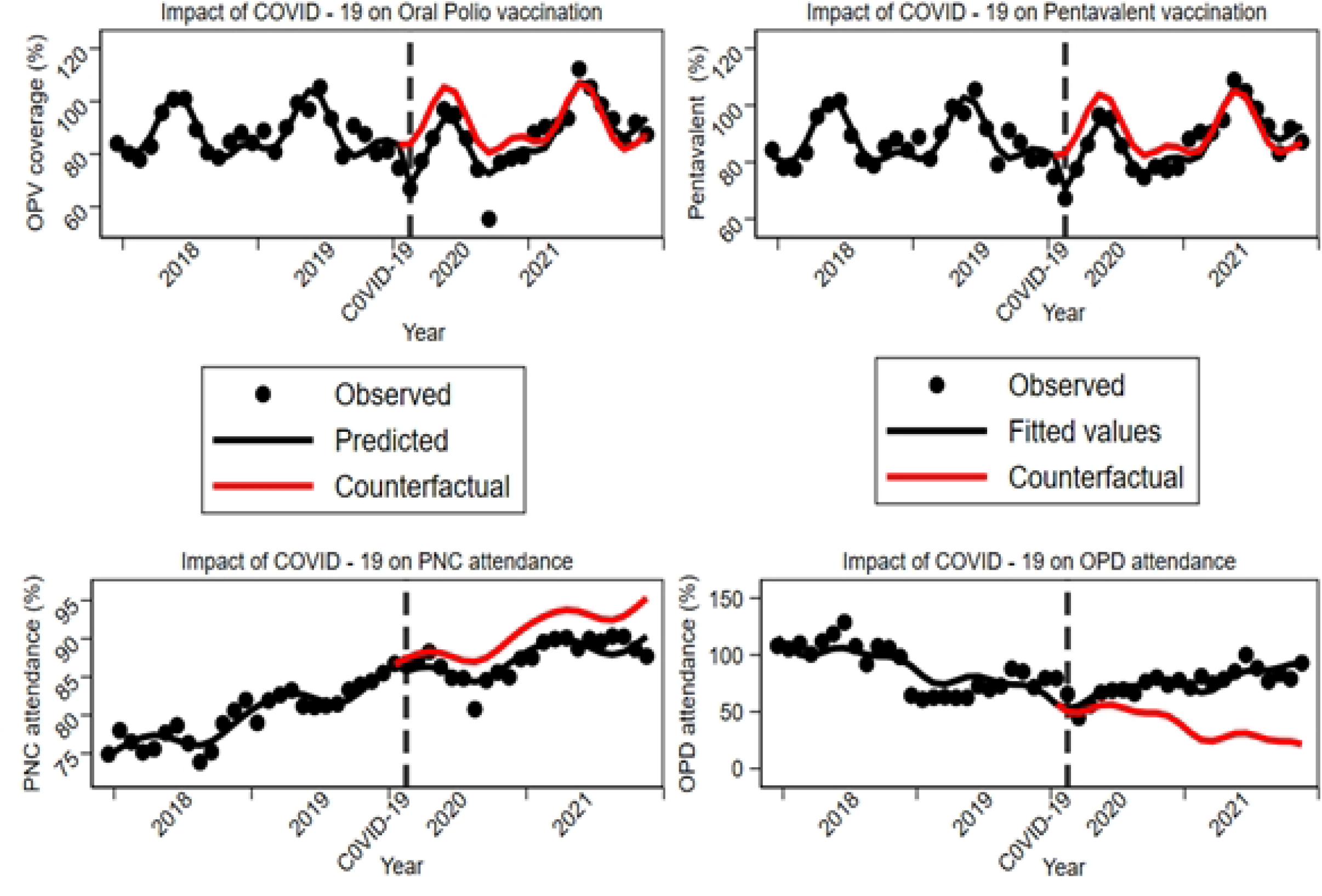
Impact of COVID-19 pandemic on the provision of essential health services.

### COVID-19 Outbreak Control Strategies

The results from the key informant interviews showed seven broad categories of strategies that were adopted to mitigate the impact of COVID-19 in Ghana: (1) rapid response coordination with preparedness & response plan leading to a reduced number of new cases and deaths: swift partial lockdown in hardest-hit regions (Greater Accra and Kumasi in the Ashanti region), (2) the adoption of pre-existing surveillance systems, leveraging previous experiences for managing epidemics; supplemented by community serological surveys provided real-time data for decision-making, (3) the implementation of 3T’s strategy: test, trace, and treat; swift contact tracing & isolation measures in early phases of the pandemic, ramp up testing through various strategies, leveraging existing GeneXpert lab equipment, pooled testing, improved vaccination strategy, private sector engagement, mass testing in hotspots, and early RDT adoption contributed to limiting the spread of the disease and preventing deaths. (4) Sustained multisectoral coordination mechanisms with political leadership commitment, including academia, donors, and private sector; mirrored at regional and district level increased the production of PPEs and. (5) the adoption of innovative technologies: use of drones to deliver COVID-19 lab samples; roll out of SORMAS for E-surveillance & reporting and (6) the employment of effective media engagement protocols with regular Presidential addresses, Ministerial press briefings, dedicated COVID-19 websites, and through social and traditional media communications. We provide a brief explanation for each exemplary practice as follows:

Theme 1: Government commitment and Rapid Response Coordination

Participants observed interventions such as private sector collaboration, legalization of non-pharmaceutical interventions, training of health personnel about the COVID-19 disease, and construction of laboratory sites.

*Sub-theme 1: Private sector collaboration*

A participant highlighted the role of private sector groups in the provision of hand sanitizers and other essential services.

> “*His Excellency the President met the various private sector groups, especially the private pharmaceutical companies, [and] within a matter of 48 hours, about 37 different companies put one of their lines to produce [hand] sanitizers. And you remember that [hand] sanitizer was a very essential commodity. Within a matter of about 2 to 3 weeks, it became something everybody could afford, and the price went down considerably*.” *(KII2)*

*Sub-theme 2: Establishment of new Laboratory Sites*

Some participants mentioned that the ability of the government to build more laboratory testing sites helped in the fight against the COVID-19 pandemic in Ghana.

> “*When we started, they were only two. [Things began to change] when the government then allowed that the private sector and the rights protection join in this fight, and, within a matter of months, we now had about 45 laboratory testing sites. More than half of them were private-owned*.” *(KII2)*

*Sub-theme 3: Training on infection prevention control*

Participants were trained on infection, prevention, and control of the COVID-19 disease.

> “*Staff commitment to training on IPC and their readiness to stick to infection prevention control measures really helped us*.” (IDI2)

> *That was when, you know, national training actually kicked started, even though some prior stakeholder engagement had taken place, but it had to take the announcement of those cases for case management training, contact tracing, surveillance, in terms of their whole response. And national*’*s trainer of training began after we had announced our cases. So, yes, it shook us, but then, I think that we began responding in doing all the things that had to be done (KII12)*

*Sub-theme 4: Non-pharmaceutical Interventions*

Participants agreed that government NPIs, backed by legal legislation, helped to control the spread of COVID-19. One of such NPIs is the closure of land, air, and sea borders.

> “*I think one important thing that we did that most countries didn*’*t do is the limitation of the importation of business by closing all our borders. Luckily, we have only one [international] airport, so [it was easy to see at first hand the cases that were being imported]*.” *(KII2)*
>
> *Sub-theme 5: Adherence to COVID-19 Protocols*

Participants indicated adherence to COVID-19 protocols by both patients and healthcare providers.

> “*At the height of the pandemic everyone was adherent to wearing masks, even though people were wearing the wrong types of masks and were wearing them in various ways when interacting with patients. But with regards to gowns, head gears, and others worn by the team [while] interacting with the patients, those were strictly adhered to in my facility*.” (IDI3)

Theme 2: Improved Disease Surveillance System

Participants explained surveillance systems such as DHIMS and SORMAS that were used for gathering accurate data during the COVID-19 pandemic.

> “*We have a platform where, on a weekly basis, all the district officers on the platform share their planned activities for the week. At the regional level too, we look at the data that we have on the DHIMS. So we identify the districts that have some challenges or need assistance and assist them. We also look at the data for priority diseases like epidemic-prone diseases [such as] yellow fever and measles. We also download the data on the DHIMS and follow up on the cases reported to verify the numbers*.” (IDI2)

Theme 3: Implementation of Testing Strategies

Testing strategies reflect different types of COVID-19 tests and preventive strategies employed by the case management team. Three sub-themes were generated from this theme.

*Sub-theme 1: Covid-19 3T (Trace, Test and Treat) Strategy*

A participant noted that healthcare providers used the 3T strategy. This involved testing a patient and when tested positive you do tracing of close relatives or neighbors while the Covid-19 patient is isolated for treatment.

> “*…[for] the testing strategy…you identify first, you test, [and] if it*’*s positive, you isolate. Then you have to do contact tracing. But the basic idea was that the moment you hear of anybody having the symptoms or anybody reporting [to have] symptoms, you quickly take their sample [and] test. When it*’*s confirmed, you go back [and] do contact tracing while the [sick] person is isolated. That was the strategy [that was] communicated to all the testing sites [and] disease control officers*.” *(KII4))*

*Sub-theme 2: Pooling*

A participant observed that one of the unique testing strategies employed by Ghana’s COVID-19 data management team was pooling. By pooling, individual samples are tested to save time and cost, as well as reduce the use of test kits.

> “*The second one was the method we call pooling. The pooling [began] when we had so many contact tracing samples. When somebody tested positive, we went and collected a lot of samples around, and, instead of working on individual samples, we pooled samples together—sometimes five samples in one or ten samples in one. So, if you have 100 samples, in the end you have only 10 to work with. So, on a daily basis, we were sometimes able to process about 5,000 samples*.” *(KII10)*

*Sub-theme 3: Vaccination strategy*

Participants identified that the government’s initial approach to vaccination was legislation on compulsory vaccination before immigrant enters the country. The policy on compulsory vaccination was eased when infection rates declined in the country.

> “*…when we started doing the vaccination, we also made sure that, if you are fully vaccinated you are allowed in. If you are a Ghanaian or resident in Ghana and you are not fully vaccinated, we give you the chance to vaccinate when you reach the airport. These are some of the things, and when things went down, we began to slowly ease the restrictions*.” *(KII2)*

Theme 4: Health System Preparedness

Participants explained the existing emergency preparedness plan used to counter any outbreak in the various facilities.

> “*The facility has an emergency preparedness plan for all eventualities and outbreaks. They have a plan, so if there is an outbreak they use that plan, and if there are some changes, they incorporate those changes*.” (IDI8)

A participant part of Ghana’s COVID-19 Taskforce explained how the government’s sustained multisectoral coordination augments the fight against COVID-19.

> “*The National Health System in place is a bit broad. At the Ministry of Health and Ghana Health Service, we have facilities at the national, regional, and district levels. At each of these levels there are facilities for prevention and case management to respond to public health emergencies. Knowing that the health system and the determinants of health are very broad, the system does not end only in the health sector. Various components and players and actors whose activities impact on public health are considered as part of the National Health System*”. *(KII7)*

> “*The Emergency Operation Center is part of the National Surveillance structure. There is now version 3 of the Integrated Disease Surveillance Response (IDSR). With regards to Public Health Emergency Management Systems, there were national-, regional- and district-level systems. At the national level, we have the National Coordination Committee, which is multi-sectoral and multidisciplinary supposed to be chaired by at least the Minister of Health and sometimes, the interior minister. The technical part is managed by the National Technical Coordinating Committee (NTCC). This is also multi-sectoral and multidisciplinary, but mostly technical in bringing together all the players and actors whose activities impact the health emergency and health sector*.” *(KII7)*.

Theme 5: Public Engagement

*Sub-theme 1: Public Education*

A participant opined that the peak stage was characterized by intense public education on the pandemic, as facilities were receiving more COVID-19 cases.

> “*At the peak stage, [public] education [intensified]. So, people*’*s fear was not as bad as it used to be, even though we were having a lot more cases in our facility*.” *(IDI18)*

### Strategies to maintain essential health services

The following practices were adopted to provide EHS: Leadership & Governance (policy guidance developed and disseminated with modified service delivery models, virtual meetings, training & patient follow-up, non-closure of health facilities), Health Care Workforce (HCW) capacity building around infection prevention control (IPC) practices for HCWs, and HCWs prioritized for vaccination, special monthly allowance for frontline HCWs and lab technicians, the suspension of service delivery for all elective medical procedures, use of an appointment system for MCNH services at the facility, effective communication, PPE availability was ensured at facilities, timely order of logistics to prevent stock out of essential supplies, medical drones were used to supply vaccines, transport blood to advance labs for testing and other essential equipment for service delivery in situations where transportation was not readily available.

*Sub-theme 1: Non-closure of facilities*

The health facilities were opened for 24 hours to allow patients to visit for any medical need at their convenience time. In addition, the public was assured that the health facilities adhere to strict screening to reduce the risk of getting infected at the health facility.

> “*As I said, we didn*’*t close any health facility. They were running 24/7 as they used to, and as part of the education that we gave the people, we told them that we were still working, so anybody who had a challenge could still come to the hospital. We even showed them our workflow, which included screening everyone entering the facility, to assure them that the risk of getting infected at the health facility was low*.” (KII1)

*Sub-theme 2: Triage Station*

Facilities instituted a station where patients were screened for Covid-19 before entering the health facilities. Establishing a triage station ensured that there was a designated physical area where frontline health workers assessed and refer patients to reduce transmission of Covid-19 and also ensured rational use of scarce resources.

> “*But we quickly also got around that by introducing measures in terms of triaging, screening patients*” (KII5)

> “*There was a triaging system put in place where, for anyone who came in, you ensured first of all that the person did not have COVID-19 before you proceeded to do anything else. COVID-19 tests were the first things we did for patients who presented*.” (IDI11)

*Sub-theme 3: Patient Appointment*

One of the innovative strategies employed by health facilities is the appointment system. By appointment strategy, healthcare professionals scheduled meeting times with patients. The appointment strategy reduced the effects of the inadequate number of frontline health workers in the facilities due to additional tasks and responsibilities resulting from COVID-19-related activities.

> “*The appointment system helped. It reduced patients*’ *wait time to see a doctor. COVID-19 taught us to use such a system and reduce the number of trips chronic patients had to make to the hospital for a refill of their medication. Our capacity in terms of testing for COVID-19 was lacking, but the appointment system helped to regulate visits to testing sites*.” (KII12)

*Sub-theme 4: Provision of PPEs*

Policymakers and implementers explained that PPEs such as face masks and goggles were distributed across health facilities in Ghana. In addition, health care workers were educated to always put on the right PPEs. This was, however, confirmed by the healthcare professionals.

> “*And then every single person in the hospital, and of course, those going out had to be in the right PPE, i*.*e*., *face mask, and where possible, goggles*.” (KII11)

*Sub-theme 5: Motivated Staff*

Policymakers and implementers explained that health personnel were encouraged to put in their best in the dire moments when the government and the people of Ghana needed them most.

> “*As leaders of the health system, we were taxed to motivate our staff to be at post, because the risk was for all. As managers, we needed to motivate our people and encourage them—not financially—whenever we could to let them know how important their role was in the management of the COVID-19 outbreak, so we did that*.” (KII11)

*Sub-theme 6: Community engagements and home visits*

Participants observed that health workers were moving into the homes of patients to provide them with health services. Home visits were a targeted approach to patients.

> “*Within the period, antenatal services were not affected much because we used to go to pregnant women in their homes*.” (KII1)

There were community engagements to encourage the local people to attend hospitals for their good.

> “*We engaged with the community to let them know that [people could come to the hospital]. Initially, the communication was that if you were not sick, you should not come to the hospital, so when things changed, we needed to go back to them to let them know that all services were back and running, and they could come*.” (KII8)

*Sub-theme 7: Redistribution and rotation of staff*

In order not to be crowded at the health facility and increase the risk of infection, health facilities were running a staff rotation (shift) system. The system allowed for few nurses in the facility at a time.

> “*I mean, we reset the minimum that was needed so that they came in batches, so that if a cohort of a batch got infected, there would be other people to continue the work*.” (KII5)

> “*As much as possible, instead of running three shifts, we decided to run two shifts: 8am to 5pm and 5pm to 8am. This was to ensure that at any point in time, there was no congestion in the hospitals*.” (KII1)

Units without adequate staff were provided with additional staff by units with more staff members. Respondent: *One of them was the redistribution of some of the staff. We got staff from units that had more staff*. (IDI1)

*Sub-theme 8: Public Education*

There was regular education by the media regarding the COVID-19 pandemic. Citizens were assured that all precautionary measures were strictly adhered to in the health facilities, hence, an assurance of minimal risk of infection.

> “*With the information we heard on radio people began to understand that no matter how we felt, we needed to report to a hospital for the hospital to decide whether it was COVID-19 or not. [It was safe] to go to the health facilities because all the precautions were observed there, and our chances of getting COVID from the health facility were quite minimal*.” (KII6)

*Sub-theme 9: Transportation Support*

Some participants were supported with the provision of buses that took them to their workplace.

> “*They made sure there was a driver and a vehicle. Any time we took a sample we called the driver to make sure they got the sample to the reference lab*.” (IDI7)

*Sub-theme 10: Telemedicine and drone services*

Some facilities employed innovative ways of providing health services to their clients. One of such innovative approaches was the use of telemedicine and drone services.

> “*We had an app, COVID Connect, by which people we treated and discharged could keep in contact with us, and people with symptoms could go into the app and indicate their symptoms for some healthcare professionals from our facility to contact them*.” (IDI11)

> *We made use of it [drones], it assisted us especially when we had very few testing centers. We were able to use the Drones to gather all the samples to the Drones Distribution center and send them to where we were having only two testing sites at Korle-Bu, Noguchi and ACCR to make use of the Drone system. And during the time that we were also doing the vaccination, we also made use of the Drone system because the Drone distribution center is like the Medical stores. (KII2)*

## Discussion

In this study, we quantified the impact of COVID-19 on the delivery of EHS, explored practices adopted to maintain the provision of EHS, and identified exemplary IPC strategies in Ghana. Our results showed that the pandemic disrupted the provision of EHS especially immunization coverage indicators within the first month of the pandemic similar to what has been reported elsewhere [14, 15]. This study highlights that with the rapid spread of COVID-19, mothers avoided taking their children to health facilities to seek child health services including immunization and vaccinations. An in-depth analysis of the results showed that during the early phase of the pandemic, there was fear of contracting COVID-19 at the health facilities, stigma, lack of innovative approach to delivering EHS, inadequate logistics such as the supply of PPE waned public trust in visiting health facilities under COVID-19 conditions, inadequate HCW for COVID-19 surge capacity proved as a challenge to the utilization of essential health services, financial challenges and low enthusiasm to work among healthcare workforce contributed to the initial disruption of EHS. This could also be attributed to imposed travel restrictions by the government of Ghana during the early days of the pandemic in March 2020 which affected the physical accessibility of EHS facilities. Findings illustrated that fear and stigma influenced the reduced health care in facilities. This aligns with the study by Al-Zaman (16) which indicated that fear and anxiety about the pandemic among communities affected the utilization of EHS. The inadequate health workforce for COVID-19 surge capacity demand proved as a challenge to the utilization of EHS.

As mentioned earlier, although COVID-19 disrupted EHS during the early phase of the pandemic especially OPV and Pentavalent vaccination among children under five, the impact was not long felt due to the systems that were put in place to maintain the provision of EHS in Ghana. These includes new policy guideline that were developed and disseminated with modified service delivery models, non-closure of health facilities to allow access at all times, use of appointment systems, provision of PPEs, staff motivation for frontline HCWs and lab technicians, home visit to provide EHS, redistribution and staff rotation to allow few nurses at the facility aimed at controlling infection and improving EHS delivery, intense public education geared towards encouraging facility visits, use of telemedicine and drone services to supply essential medical equipment’s in situations where transportation was not readily available, capacity building around IPC practices for HCWs and the prioritization of HCWs for early vaccination to reduce the fear of patients getting infection at the facilities, service delivery suspended all elective medical procedures to focus on providing other EHS and treatment of COVID-19 related cases, use of an appointment system for MCNH services at the facility, effective government communication and commitment to increase access to EHS during the peak phase of the pandemic that resulted in-home visits, and the timely order of logistics to prevent stock out of essential supplies. Literature highlights population-based intervention at interpersonal, institutional, community and public policy levels to sustain the delivery of EHS. At the interpersonal level of intervention, positive COVID-19 attitudes and perceptions were promoted through public education and routine home visits.

As a form of institutional intervention, the study finds that health professionals were reassigned to other units with inadequate staff. This inadvertently led to a reduction in the utilization of EHS such as child health services as found in other studies [17]. With the upsurge of Covid-19, building a resilient health system in SSA was a challenge. We found that one of the institutional interventions implemented to reorganize and maintain access to safe and EHS was the screening and establishment of triage in health facilities. The screening and creation of triage of suspected COVID-19 patients reassured them of their safety in the health facilities when they visit. Another institutional intervention used was telemedicine. Using telemedicine approaches such as COVID Connect – an application that keeps patients and health workers in contact proved critical to providing EHS. Among other benefits, this application identifies hotspots, manages and registers attendance at events, tracks government updates and monitors symptoms. Other studies indicated that telemedicine increases trust, and fosters feelings of intimacy and relief when health workers and the patient already know each other [18, 19].

At the public policy level, the study found that the government of Ghana implemented different intervention packages to motivate and protect frontline health workers at the forefront of the pandemic fight. The packages include providing them with an allowance of 50% of their basic salary and also making available free buses to and from work at specific routes. Other interventions at the public level include non-closure of facilities and provision of PPEs in health facilities for frontline health workers. This assured the public of availability and readiness to serve their health needs. The government also distributed PPEs across the country to increase the safety of frontline health workers and patients.

There is a correlation between IPC strategies and the provision of EHS. Adopting effective prevention control measures reduces the burden of the pandemic on health systems paving the way to provide EHS. The integrated interventions implemented by the Government of Ghana and other stakeholders contributed significantly to the minimal impact of the pandemic on the health system and this consequently increased the ability of the health system to provide EHS in the hardest hit regions.

## Strengths and Limitations of the study

This remains the only study that has simultaneously explored the complex interplay between the infection outbreak control strategies, the impact of the pandemic on EHS and practices geared towards maintaining the provision of EHS during the early phase of the pandemic. That notwithstanding, the study has some limitations. The qualitative data may not be representative of the target population as it largely reflects the opinion of the key informants that were interviewed. That is, caution and contextual consideration must be applied when the intent is to directly transfer the knowledge gained and entire results to other contexts and settings. Although more rigorous statistical methods have been employed to quantify the impact of the pandemic on EHS delivery, our study could suffer from potential selection bias due to the quasi-experimental design and caution must be applied when the intention is to infer causality.

## Conclusion

This study provides evidence of the impact of COVID-19 on the provision of EHS and exemplary outbreak control strategies. Although EHS were disrupted during the early phase of the pandemic, the complex integrated approach that includes infection prevention control strategies, resilient health system, government commitment, private sector and stakeholder engagements contributed significantly to the effective management of the pandemic and the provision of EHS.

## Data Availability

Data cannot be shared publicly because of ethical consideration but data would be release after permission from Ghana Health Service Ethical Review Committee

## Acknowledgments

We acknowledge all participants that willingly agreed and provided written or verbal consent to be part of the study.

## Notes

### Competing Interest Statement

The authors have declared no competing interest.

### Funding Statement

Financial support for this study was obtained from the Bill and Melinda Gates Foundation (INV-019313) in conjunction with Gates Ventures. The views, opinions, and content of this publication are those of the authors and do not necessarily reflect the views, opinions, or policies of the Bill and Melinda Gates Foundation or Gates Ventures. The funders had no role in study design, data collection and analysis, decision to publish, or preparation of the manuscript

### Author Declarations

Ethical approval for the study was granted by the Ghana Health Service Ethics Review Committee (GHS-ERC: 008/11/12).

